# Reinforcement learning optimization of automated insulin delivery in type 1 and type 2 diabetes mellitus

**DOI:** 10.1101/2025.10.12.25337835

**Authors:** Nelida E. Lopez-Palau, Pablo Naranjo-Meneses, Julia Szendroedi, Roland Eils, Stefan M. Kallenberger

**Affiliations:** Health Data Science Unit, University Hospital Heidelberg and Center for Quantitative Analysis of Molecular and Cellular Biosystems (BioQuant), University of Heidelberg, Heidelberg, Germany; Department for Endocrinology, Diabetology, Metabolic Diseases and Clinical Chemistry, University Hospital Heidelberg, Heidelberg, Germany; Joint Heidelberg-IDC Translational Diabetes Program, Internal Medicine, Heidelberg University Hospital, Heidelberg, Germany; Center of Digital Health, Berlin Institute of Health at Charité - Universitätsmedizin Berlin, Berlin, Germany; National Center for Tumor Diseases, Department of Medical Oncology, Heidelberg University Hospital, Heidelberg, Germany

## Abstract

Closed-loop insulin delivery systems have proven effective in regulating blood glucose (BG) concentration, thereby reducing the burden of self-care in the management of type 1 and type 2 diabetes mellitus. However, the prevalence of unexpected disturbances resulting from oral glucose intake represents a considerable challenge to the full automation of these systems. Here, we propose an actor-critic reinforcement learning (RL) framework implemented within environments governed by compartmental ordinary differential equation models of the glucose–insulin–glucagon–incretins dynamics. This approach was employed to optimize automated insulin delivery in virtual patients with type 1 and type 2 diabetes mellitus, under scenarios involving unforeseen BG disturbances. The resulting optimal RL policies were tested *in silico* on virtual patients subjected to three unannounced glucose disturbances over the course of a day. The findings demonstrated that optimal RL policies could sustain the BG a significantly higher percentage of time within the normoglycemic range and a significantly lower percentage of time below the normoglycemic range in comparison to either continuous or discrete proportional-integral-derivative control algorithms. These results set the basis for developing new approaches to optimizing automated dosing regimens for chronic disease management.

**Author Summary:** Managing diabetes requires constant attention to glucose levels and the corresponding adjustment of insulin doses, which can be demanding for insulin-dependent patients. Semi-automated systems, frequently referred to as “artificial pancreas”, aim to mitigate this burden by adjusting insulin delivery based on glucose levels obtained from a continuous glucose sensor. However, these systems underperform in scenarios involving unexpected glucose disturbances, such as those triggered by the omission of meal announcing within the system interface. In the present study, we developed a computer-based learning approach to identify deep network-based functions that determine the appropriate insulin doses to regulate glucose in real time and mitigate unannounced disturbances. This approach has been demonstrated to be effective in the management of Type 1 and Type 2 diabetes and it does not require any additional input other than the continuous glucose measurements. The findings of our study demonstrate that our functions were able to maintain glucose levels within a healthy range for a longer period of time than the standard functions, while also reducing the risk of threating low glucose levels. This research contributes to the development of fully automated insulin delivery systems that are capable of adapting to real-life situations of individuals living with diabetes.

## Introduction

Maintaining blood glucose (BG) concentration within the optimal range in individuals with diabetes mellitus (DM) is of paramount importance to reduce the risk for concomitant and secondary diseases such as neuro- or nephropathy, and minimize the incidence of severe and potentially life-threatening medical complications [1]. Accordingly, therapeutic strategies must assure robustness in BG regulation. In patients with type 1 diabetes mellitus (T1DM) the body is unable to produce insulin, thus resulting in impaired glucose transportation from the bloodstream to peripheral cells and elevated BG. Therefore, the administration of exogenous insulin is required [2]. In contrast, patients with type 2 diabetes mellitus (T2DM) are characterized by insufficient insulin availability, resulting either from inadequate secretion or from insulin resistance, that is, an impaired responsiveness of target tissues to insulin. Hence, therapy with exogenous insulin becomes imperative for individuals with T2DM who are unable to achieve adequate BG regulation through lifestyle modifications and the administration of non-insulin hypoglycemic agents [2]. In patients diagnosed with either T1DM or T2DM, the insulin strategy requires constant monitoring of the BG concentration and the continuous adjustment of insulin doses in accordance with dietary intake, the level of physical activity, tissue demands for glucose, and other factors such as stress or illness [2]. Consequently, achieving optimal insulin administration strategies in traditional DM management (*i*.*e*., self-care) entails a considerable mental burden on individuals with DM [3].

The development of automated insulin delivery (AID) systems, also called closed loop insulin delivery systems, represents a substantial advance in the field of DM care, as they aim to reduce the patient’s self-care burden for insulin dosing decisions and optimize BG regulation [4]. The underlying principle of an AID system is to calculate and continuously deliver insulin with minimal or no patient input, thereby allowing greater independence and reducing the need for constant intervention in insulin management. The AID design is based on the integration of a continuous glucose monitor (CGM) and an insulin pump mediated by a control algorithm. The fundamental function of this algorithm is to automatically adjust insulin delivery in response to BG measurements to prevent hyper or hypoglycemia. The control algorithm is therefore crucial for BG regulation not only in presence of disturbances related to meals or physical activity, but also due to inter- and intra-patient variability in insulin sensitivity and insulin absorption as well as delays in the glucose and insulin response [5].

In the last decades, considerable efforts have been made to develop effective control algorithms for insulin dosage in AID systems. Early efforts in this area included linear approaches derived from classical control techniques, with Proportional-Integral-Derivative (PID) control being a prominent example. The PID controller can regulate BG by adjusting a limited set of parameters to proportionally respond to instantaneous errors between target and measured glucose values, integrate the cumulative error over time, and account for the derivative of the error signal [6]. Despite their simplicity and accessibility, PID control algorithms, as well as other linear techniques, are unable to adequately address the uncertainty and inherent nonlinear dynamics of glucose metabolism [7]. In response to these limitations, researchers have developed nonlinear techniques, for instance model predictive control (MPC) [8], backstepping control [9], or fuzzy logic [10] to enhance the accuracy and precision of control strategies. A comprehensive overview of the most prevalent control systems to regulate BG is provided by Mughal, e*t al*. [11].

Since the approval of the first AID system for BG management by the United States Food and Drug Administration in 2017 [12], the predominant control systems in commercial devices are based on classical control algorithms, namely PID or MPC [13]. Due to their simplicity and stability, AID systems, such as *MiniMed* devices, commonly employ PID control algorithms to regulate BG in patients with T1DM [14]. Nevertheless, managing BG fluctuations associated with meals remains challenging.

Recently, optimized learning algorithms enabled the implementation of reinforcement learning (RL) for regulation of BG [15] with strong emphasis in T1DM [16]. RL is a goal-directed approach, in which an agent interacts with an environment in order to explore and learn the optimal way of performing a task without human intervention (*i*.*e*., policy) [17]. In the context of BG regulation, this adaptive learning capability has enabled the development of control algorithms designed to respond to postprandial BG fluctuations. A common learning strategy involves offline methods, where the policy is adapted using historical data, *e*.*g*., medical records [18–20], or previous documented BG features [21–23]. Although this approach provides stability and safety during training, its major limitation is the need for a large amount of training data [19,21]. To overcome this limitation, online methods have been proposed as an alternative, adapting the policy in real time as the new patient data is received, without relying on historical data.

To date, some online RL algorithms have been proposed to regulate fasting and postprandial BG in T1DM, implementing a variety of training methods, including Q-learning [24,25], soft actor-critic [26], and proximal policy optimization (PPO) [27,28]. A limitation of these control algorithms is their reliance on manual patient input concerning meal intention (*i*.*e*., meal announcement) [27] and estimation of meal content (*i*.*e*., carbohydrate counting) [25,26,29]. It is noteworthy that specialized training and expertise are prerequisites for accurate carbohydrate counting, which frequently results in imprecise estimations and inaccurate BG regulation [30]. In the context of T2DM, a normalized advantage function in conjunction with a minimal model has been implemented to automate insulin delivery without requiring meal announcement or carbohydrate counting [31]. Notwithstanding the encouraging outcomes, the minimal model of the latter study did not consider BG measurement disturbances, intra-patient variability, or other complexities inherent to the glucose-insulin dynamics. In this perspective, it can be anticipated that in the future, AI-driven applications in the therapy of T1DM and T2DM patients can profit from further generalization to pathophysiological aspects and heterogeneity in patient populations to support their translation to the clinics.

In this study, an online RL approach in combination with ordinary differential equation (ODE) models of glucose-insulin-glucagon-incretins dynamics was implemented to determine the rate of insulin delivery required to compensate for BG fluctuations resulting from unannounced oral glucose intakes and other common BG disturbances. To accomplish this objective, a PPO method was employed to train two RL agents, one for regulating BG in T1DM patients and another targeting T2DM patients. During training, the agents interacted with virtual patients (VP) described by ODE models related to the pathophysiology of either T1DM or T2DM. As a result, optimal RL policies were obtained, requiring only CGM measurements to compensate for unannounced oral glucose disturbances, thereby obviating the need to specify, infer or predict the amount of ingested glucose or to announce meals. The efficacy of the optimal RL policies was evaluated against PID control algorithms, selected as the standard benchmark for comparing novel BG regulation systems. Ultimately, these optimal RL policies could be implemented as control algorithms in AID systems to regulate BG fluctuations in patients with T1DM and T2DM, eliminating the need for user intervention.

## Results

### Reinforcement learning-based framework for insulin delivery control in blood glucose regulation

In order to optimize the regulation of undesired BG fluctuations in T1DM and T2DM, we developed a closed-loop control approach, integrating deep RL and compartmental mathematical modelling of BG dynamics. In this approach, BG regulation was resembled by a sequence of interactions between an agent and its environment (**Fig 1a**). The reinforcement learning (RL) agent, implemented using artificial neural networks (ANNs), was integrated with the insulin delivery and control systems, while the environment was associated with the BG dynamics observed through the measurements of a CGM over time. The interactions were conducted in episodes, subdivided into a series of steps. At the initiation of each step, the state of the system *s*_*t*_ ∈ *S* was observed and transferred to the agent. The state contained information derived only from CGM measurements obtained along the step time-lapse every *T*_1_ min (*i*.*e*., observations), for instance, the rate of change of BG (*dBG*_*m*_), the average of the BG (*BG*_*m*_), the discrepancy between the BG and the normoglycemic reference (*BG*_*e*_), and an attenuation factor (*φ*), which forced the immediate cessation of insulin delivery in case the BG approached a hypoglycemic range and a downward trend was detected. Based on the current state, the agent autonomously selected an action *a*_*t*_ ∈ *A* ⊂ ℝ to be conducted by employing the policy *π*. The action constituted the rate of continuous insulin delivery in the current step. At the end of each step, the environment evolved into a subsequent state, and the consequence of the action was assessed by a delayed reward (*r*_*t*+1_ ∈ *R*⊂ℝ). The reward function quantified the effectiveness of each action, encouraging rapid attainment and maintenance of BG within the target range (*i*.*e*., 90 to 120 mg/dL, **Fig 1b**) while discouraging hypoglycemic states (see Methods for details).

**Fig 1.**
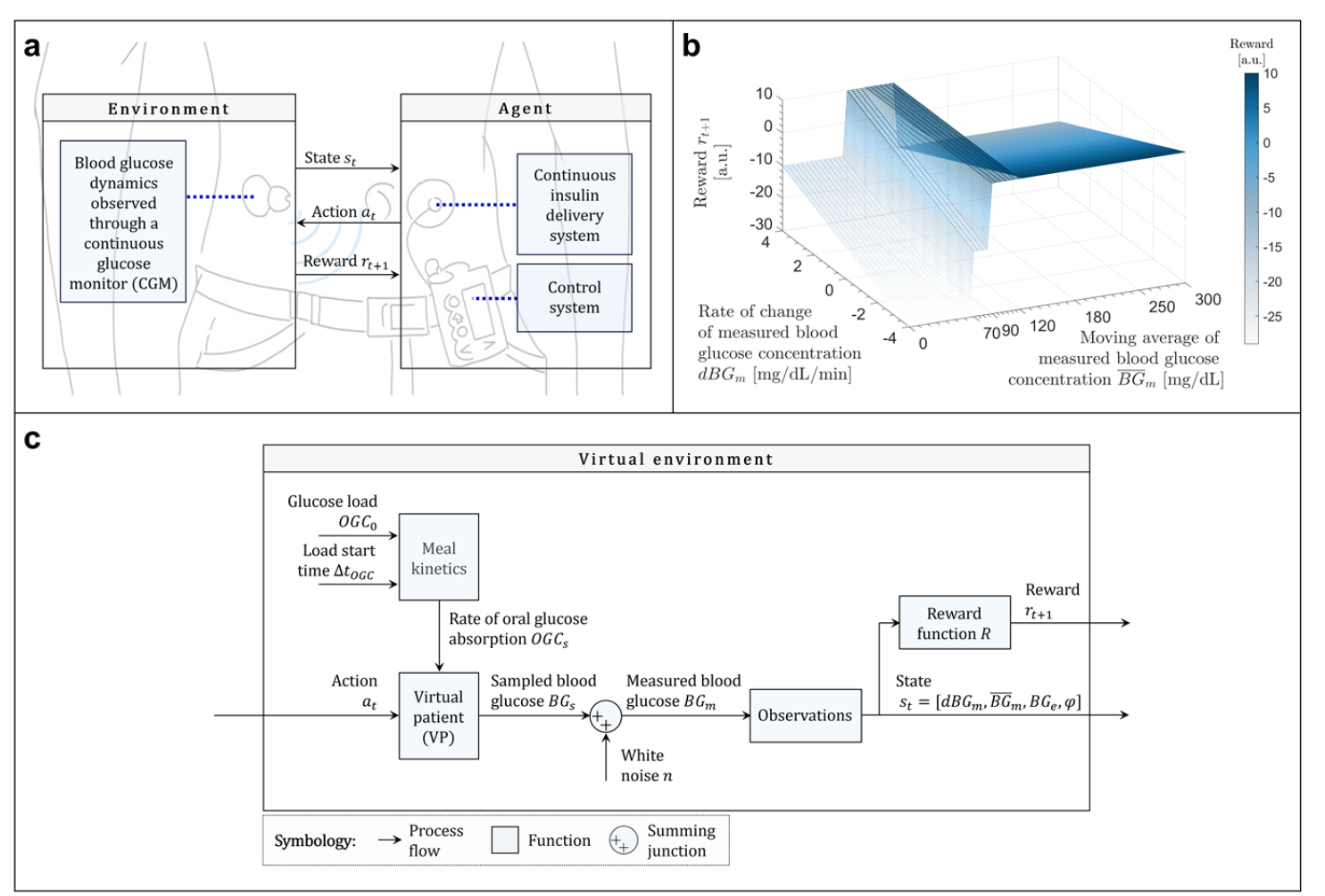
Closed-loop control of blood glucose concentration using a reinforcement learning approach. (**a**) This study investigated algorithms of automated insulin delivery systems for regulating BG in DM patients. In the context of RL, the control system used an optimal RL policy to adjust insulin release (actions) in response to the state derived from continuous BG measurements (environment). BG regulation was achieved through interactions in a cycle of actions, states and delayed rewards (represented by arrows). (**b**) The reward function was dependent on the rate of BG change and the average of the BG during the current step. A high reward was assigned when BG was within a targeted range (90 to 120 mg/dl). Conversely, negative values reflected penalties for hypoglycemic states (below 70mg/dl). (**c**) The block diagram illustrates the virtual environment implemented for the training and validation of RL agents. The arrows represent the flow of information, emphasizing processing of actions, the effect of disturbances, and the generation of states and rewards through measurements of the BG.

The policies to regulate BG in T1DM and T2DM patients, hereafter referred to as *π*_*T*1*DM*_ and *π*_*T*2*DM*_, respectively, were optimized by training in a virtual environment (**Fig 1c**). In the virtual environment, BG values were obtained from VPs implemented by simulating ordinary differential equation (ODE) models.

These ODE models described compartments, resembling organs of the human body interconnected by blood flow. Each organ was further subdivided into sub-compartments to consider flow exchange between vascular, interstitial or intracellular spaces. The changes in glucose, insulin, glucagon and incretin concentrations in sub-compartments were simulated by mass balances (see **S1 Text** and **S1-S6 Tables** for details). T1DM and T2DM VPs were simulated using distinct model parameterizations. For instance, the T2DM model incorporated a residual rate of endogenous insulin secretion, which is not considered in the T1DM model (see **S1 Text**). To account for the heterogeneity of the patient population, a subset of parameters from each VP (*e*.*g*., peripheral fasting glucose and insulin concentrations, or oral glucose dose) was sampled from distributions (see Methods section for details). Subsequently, a sample of VPs was selected for the training and validation environments.

For training, the policies were initiated with arbitrary initial parameters *θ*_0_ and defined as 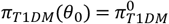 and 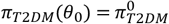. Subsequently, the policies were optimized by interacting with the training environment over multiple episodes. Each training episode *e* lasted 3 hours and consisted of *N* = 12 time steps of *T*_2_ = 15 minutes with BG measurements of *T*_1_ = 5 min, in which the rate of insulin delivery was controlled. Along each training episode, the BG dynamics was disturbed by external factors included variations in the oral glucose load (*OGC*_0_), the time of ingestion (Δ*t*_*OGC*_), exogenous insulin delivery (*i*.*e*., actions), and white noise *n* affecting the *BG*_*s*_ values of the VPs. The training process entailed the identification of the set of probabilities associated with the execution of specific actions, given the state of the system at each step. The objective of this iterative process was to ascertain the set of optimal parameters *θ*\*, for which the policy systematically selected actions maximize the discounted accumulated total reward *r*_*total*_ during an episode. Optimal RL policies were denoted as 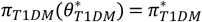 and 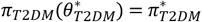.

### Validation of the optimal RL policies for T1DM and T2DM

Following the training phase, the optimal RL policies *π*\* were deployed to the agents and evaluated through simulations in both the training and validation environments. Subsequently, the cumulative rewards obtained by the optimal RL policies *π*\* in the training environment were compared to those achieved by the initial policies *π*^0^ under identical simulation conditions (**Fig 2**). Convergence of the learning process was evidenced by a reduction in the standard deviation of cumulative rewards obtained with the optimal RL policies *π*\*, relative to those observed with the initial policies *π*^0^. In T1DM as well as T2DM, discrepancies between standard deviations of the optimal RL policies *π*\* in the training and validation environments were negligible, thereby suggesting an absence of overtraining.

**Fig 2.**
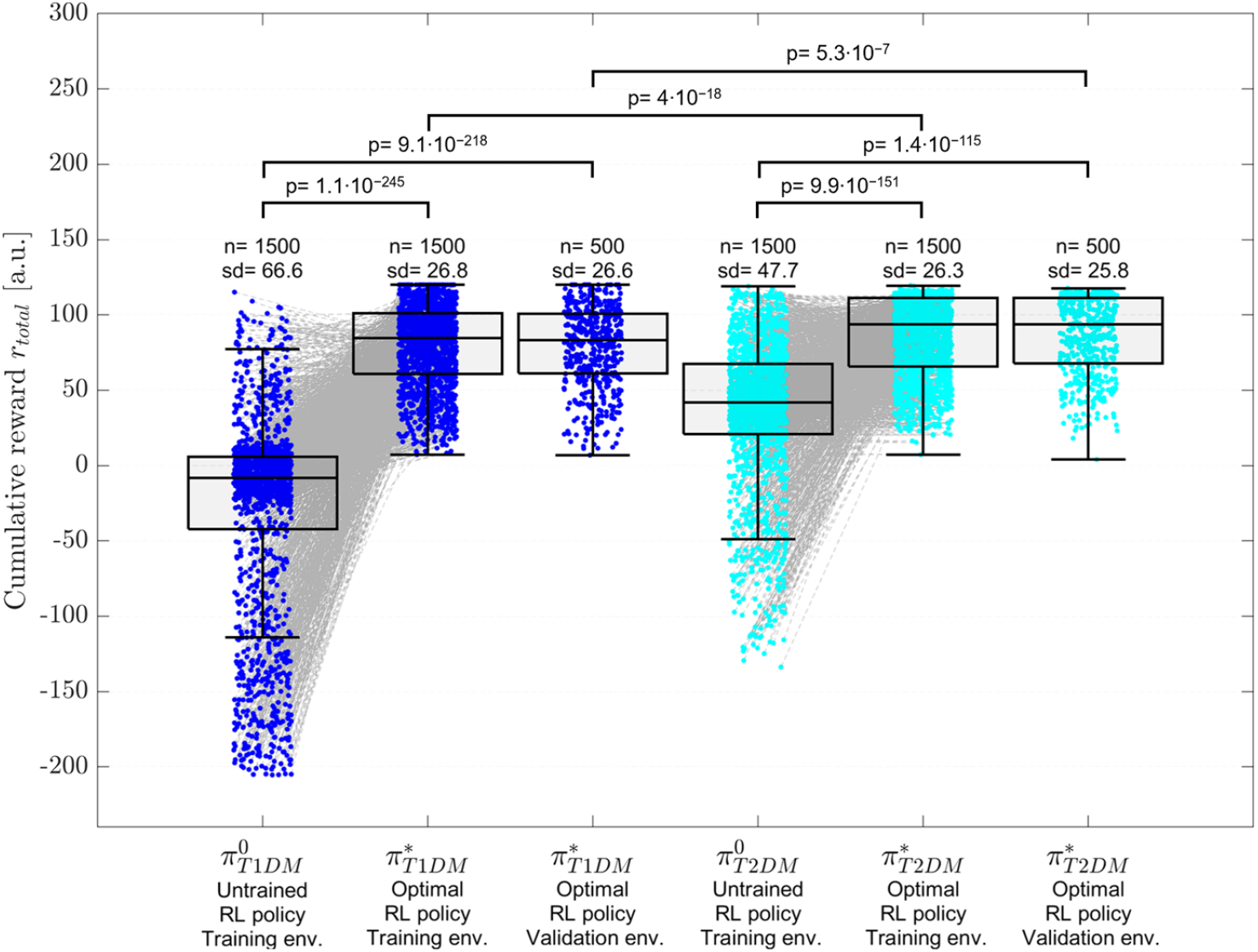
Cumulative rewards from untrained and trained policies in T1DM and T2DM. The first boxplot illustrates the cumulative reward values achieved by the T1DM initial (untrained) policy, evaluated in the training environment of T1DM VPs (n=1500). Individual simulation outcomes are overlaid as scatter points. The second boxplot presents the performance of the optimal (trained) RL policy in the training environment. Gray dashed lines connect simulations of identical VPs in untrained and trained policies. The third boxplot depicts the cumulative rewards achieved by the optimal RL policy for T1DM on the validation environment (n=500). The remaining three boxplots represent the analogous boxplots for simulations performed with T2DM VPs. Statistical significance is shown based on Wilcoxon rank-sum test followed by Bonferroni correction for multiple testing.

Simulations with the initial policy 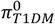 resulted in a higher frequency of adverse outcomes compared to those using the initial policy 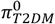. Similarly, a comparative analysis of the mean and standard deviation of cumulative rewards indicates that 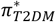 significantly outperformed 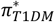 in both the training and validation environments (p = 4 · 10^−18^ and p = 5.3 · 10^−7^, Wilcoxon rank-sum test, **Fig 2**). These results can be partially attributed to two key factors. First, the T2DM VPs exhibited partially compensated BG dynamics due to residual endogenous insulin secretion, which was absent in the T1DM VPs. Second, differences in the initial model conditions contributed to the observed disparity. Specifically, based on the distributions used to define the postabsorptive basal state, T1DM VPs started the episode simulation in a broader range of initial BG concentrations relative to T2DM VPs, thereby increasing the likelihood of hypoglycemic or hyperglycemic events.

Compared to the corresponding initial policies 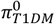, the cumulative rewards obtained with the optimal RL policies 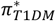 increased significantly in both the test and validation environments (p = 1.1 · 10^−243^ and p = 9.1 · 10^−218^, Wilcoxon rank-sum test). A similar improvement was observed for the optimal RL policies in T2DM (p = 9.9 · 10^−151^ and p = 1.4 · 10^−115^, Wilcoxon rank-sum test).

In a small subset of VPs, both in T1DM and T2DM, the cumulative reward was higher under the initial policies *π*^0^ than under the optimal RL policies *π*\*. Analyzing these cases revealed that, in some instances, although the optimal RL policies successfully achieved and maintained normoglycemia, the initial policies *π*^0^ could obtain a temporarily higher rewards by aggressively lowering BG concentrations, thus capitalizing on the immediate gains associated with rapid BG reduction (Eq. 5). However, when the simulation time was extended, this aggressive strategy frequently resulted in hypoglycemic episodes, which are heavily penalized (Eq. 6). Consequently, although an untrained aggressive policy could yield a short-term increase in reward, the optimal RL policy adopted a more conservative strategy minimizing the risk of costly penalizations in the long term.

The optimal RL policies 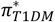 and 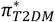 consistently achieved the highest reward value across multiple simulations in training and validation environments, notwithstanding the unannounced oral glucose disturbances and the presence of noise in the CGM sensor measurements. This robust performance indicates that the agents were able to learn an effective policy capable of generalizing to environmental uncertainty. The agents were not only able to adapt to stochastic variations, but also to mitigate the effects of uncertainties on their decision-making ability.

### Reinforcement learning-based insulin delivery outperforms PID control algorithms

Following the validation process, the optimal RL policies *π*\* were tested in a virtual environment, defined as ‘test environment’. The test environment comprised simulations involving three unannounced oral glucose disturbances over the course of a day sampled from uniform distributions (see Methods for details). Each episode in the test environment contained *N* = 96 steps of *T*_2_ minutes equivalent to one day. To test the policies in controlling postprandial BG variations, variations, three unannounced oral glucose disturbances of varying magnitudes were simulated at random times during each episode. The outcomes of the simulations with the optimal RL policies *π*\* were compared to those simulations under analogous conditions with either a continuous PID control algorithm (*PID*) or a digital PID control algorithm (*PID*^*D*^).

**Fig 3** shows the simulations conducted in the test environment for exemplary T1DM (**Figs 3a and 3b)** and T2DM VPs (**Figs 3c and 3d**). From an initial condition of fasting hyperglycemia (**Figs 3a and 3c**), the optimal RL policies *π*\* and the control algorithms *PID* and *PID*^*D*^ exhibited similar initial responses characterized by high insulin infusion rates (**Figs 3b and 3d**). Nevertheless, the optimal RL policies *π*\* achieved more stable regulation of BG within the normoglycemic range (*i*.*e*., 80 to 110 mg/dL) than either *PID* or *PID*^*D*^. Following oral glucose disturbances, all controllers regulated postprandial hyperglycemia by increasing insulin delivery rates. Both PID strategies exhibited a more aggressive approach to BG regulation during fasting and postprandial phases more frequently resulting in BG falling below the normoglycemic range (*i*.*e*., <80 mg/dL). In contrast, the optimal RL policies prevented hypoglycemia more effectively. The minor delay between the glucose ingestion and the subsequent increase in the insulin delivery rate, observed in the optimal RL policies responses, indicates that the policies did not predict the timing of glucose intake. Rather, they inferred information from fluctuations in BG indicating absence of overtraining.

**Fig 3.**
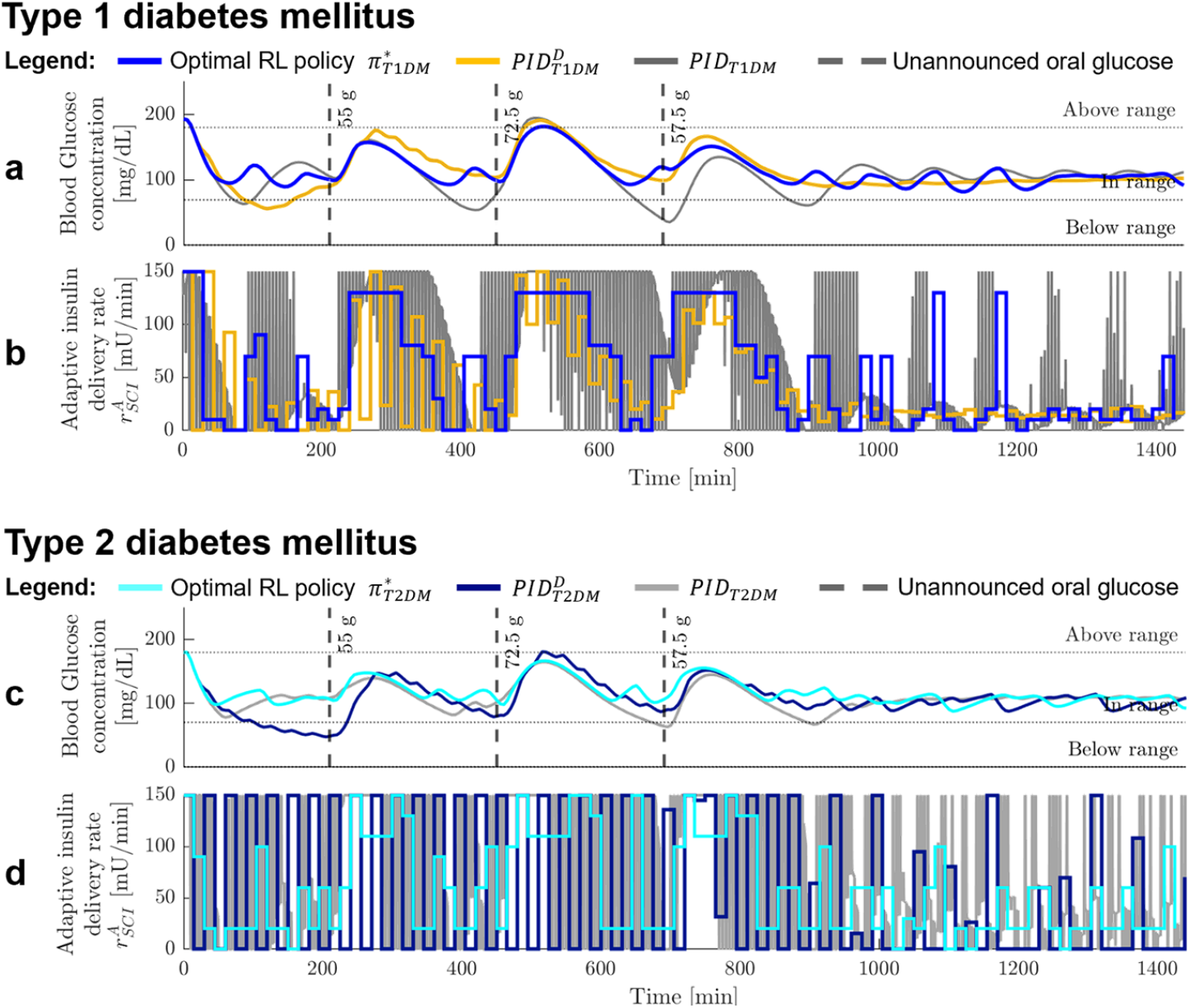
Improved BG regulation achieved by optimal RL policies compared with PID control in response to oral glucose disturbances. Panels illustrate BG trajectories and insulin delivery profiles in VPs with T1DM (**a, b**) and T2DM (**c, d**), under identical simulation conditions with matched oral glucose disturbances. Outcomes are shown for the optimal RL policies *π*\*, the *PID* controller, and the *PI D*^*D*^ algorithm. Panels **a** and **c** depict BG dynamics, whereas panels **b** and **d** present the corresponding insulin infusion rates. In these representative simulations, the optimal RL policies *π*\* demonstrated superior regulation of both fasting and postprandial BG relative to PID-based strategies, more effectively mitigating both hypoglycemia and hyperglycemia.

To compare control algorithms, one episode was simulated for n=200 VPs, each characterized by a distinct pattern of BG disturbances. The effectiveness of the control algorithms was assessed by means and coefficients of variation (CV) of the BG concentration, as well as percentages of time in range (TIR), time below range (TBR), and the time above range (TAR), see **Table 1 and Fig 4**. Mean BG concentrations of optimal RL policies *π*\* were similar to PID control algorithms (ranging between 103 and 114mg/dl). However, in case of T1DM and T2DM, the BG variability of the RL policies was lower as compared to PID controllers, indicated by smaller CV values. In general, the control algorithms for T1DM patients showed higher BG variability than their counterparts used for T2DM patients, which was particularly evident in *PID*_*T*1*DM*_. Consistent with the observations in training and validation environments, the smaller BG variability observed in T2DM within the testing environment may be attributed to differences in the distributions used to define the postabsorptive basal state for T1DM and T2DM. Overall, the performance of all control algorithms complied with the recommended threshold of CV ≤ 36%, as outlined in current clinical guidelines [32,33]. Notably, the optimal RL policy 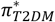 exhibited the lowest BG variability, with a CV of 10.6%.

**Table 1.**
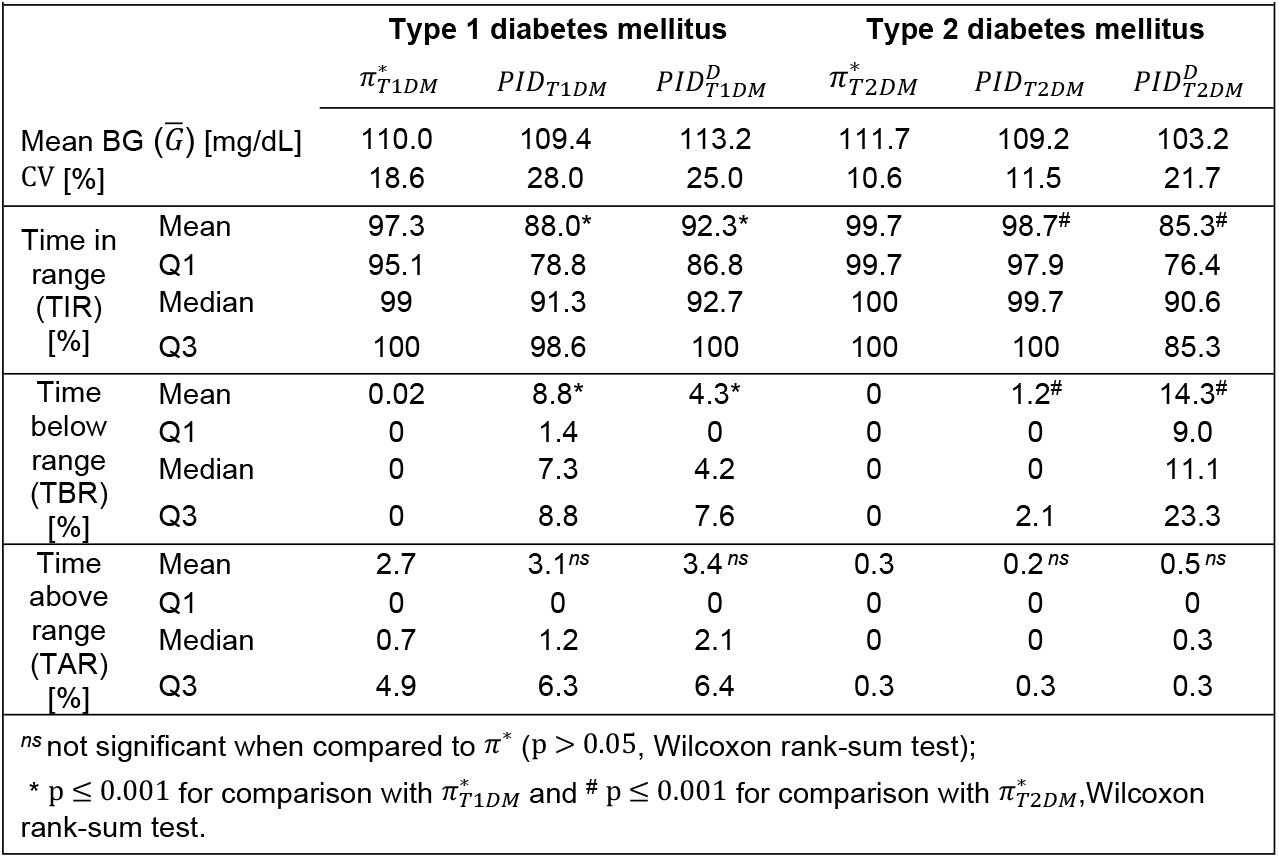
Comparison of BG regulation by optimal RL policies and PID controllers.

|  |  |  |  |  |  |  |  |
| --- | --- | --- | --- | --- | --- | --- | --- |
|  |  | Type 1 diabetes mellitus |  |  | Type 2 diabetes mellitus |  |  |
| | | $\pi_{T1DM}^*$ | $PID_{T1DM}$ | $PID_{T1DM}^D$ | $\pi_{T2DM}^*$ | $PID_{T2DM}$ | $PID_{T2DM}^D$ |
| Mean BG ($\bar{G}$) [mg/dL] | | 110.0 | 109.4 | 113.2 | 111.7 | 109.2 | 103.2 |
| CV [%] |  | 18.6 | 28.0 | 25.0 | 10.6 | 11.5 | 21.7 |
| Time in range (TIR) [%] | Mean | 97.3 | 88.0* | 92.3* | 99.7 | 98.7<sup>#</sup> | 85.3<sup>#</sup> |
|  | Q1 | 95.1 | 78.8 | 86.8 | 99.7 | 97.9 | 76.4 |
|  | Median | 99 | 91.3 | 92.7 | 100 | 99.7 | 90.6 |
|  | Q3 | 100 | 98.6 | 100 | 100 | 100 | 85.3 |
| Time below range (TBR) [%] | Mean | 0.02 | 8.8* | 4.3* | 0 | 1.2<sup>#</sup> | 14.3<sup>#</sup> |
|  | Q1 | 0 | 1.4 | 0 | 0 | 0 | 9.0 |
|  | Median | 0 | 7.3 | 4.2 | 0 | 0 | 11.1 |
|  | Q3 | 0 | 8.8 | 7.6 | 0 | 2.1 | 23.3 |
| Time above range (TAR) [%] | Mean | 2.7 | 3.1<sup>ns</sup> | 3.4<sup>ns</sup> | 0.3 | 0.2<sup>ns</sup> | 0.5<sup>ns</sup> |
|  | Q1 | 0 | 0 | 0 | 0 | 0 | 0 |
|  | Median | 0.7 | 1.2 | 2.1 | 0 | 0 | 0.3 |
|  | Q3 | 4.9 | 6.3 | 6.4 | 0.3 | 0.3 | 0.3 |
| <sup>ns</sup> not significant when compared to $\pi^*$ ($p > 0.05$, Wilcoxon rank-sum test);<br>* $p \leq 0.001$ for comparison with $\pi_{T1DM}^*$ and <sup>#</sup> $p \leq 0.001$ for comparison with $\pi_{T2DM}^*$, Wilcoxon rank-sum test. | | | | | | | |

**Fig 4.**
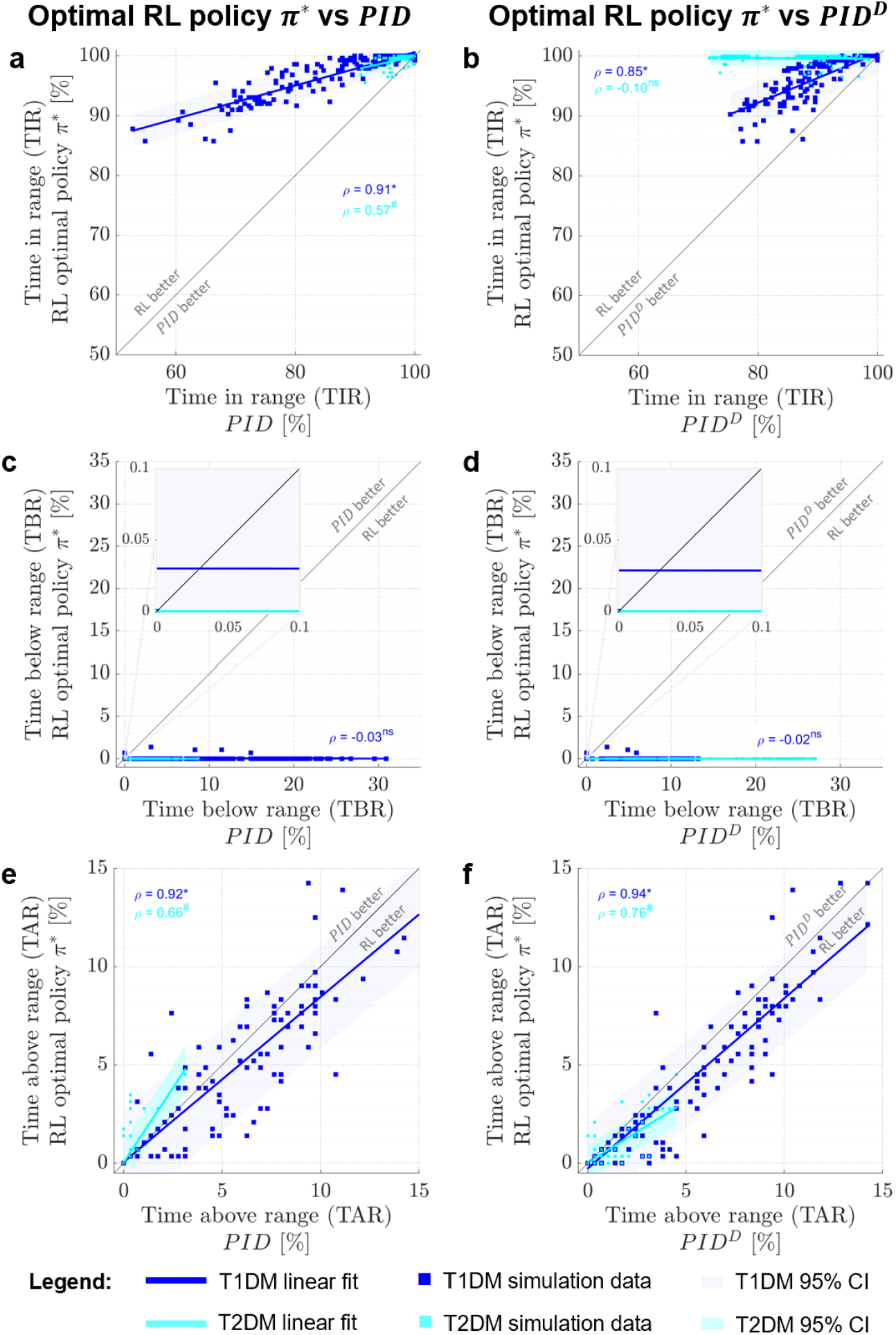
Comparison of performance indices between optimal RL policies and PID control. Left panels show comparisons between the optimal RL policies *π*\* and the *PID* control of time spent in range (**a**), below range (**c**), and above range (**e**). Right panels (**b, d**, and **f)** display the corresponding comparisons between the optimal RL policies *π*\* and the *PID*^*D*^ control. Each point represents the percentage of time in glycemic ranges for the n=200 VPs across one-day episode simulations in the test environment. The correlation between the control strategies has been evidenced by linear regression [ρ, Pearson correlation coefficient; shaded areas, 95% coefficient interval (CI) of linear fits; *, p ≤ 0.05 for correlation with 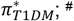, p ≤ 0.05 for correlation with 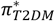; ns, p > 0.05; p-values obtained using a Student’s t distribution for transformation of the correlation].

The effectiveness of the long-term BG regulation was assessed by the TIR percentage. The analysis in T1DM and T2DM patients consistently demonstrated higher TIR percentages for the optimal RL policies as compared to both PID control strategies (**Figs 4a and 4b**). In T1DM VPs, the optimal RL policy 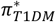 achieved significantly larger TIR percentages than both *PID*_*T*1*DM*_ (p = 1.4 · 10^−16^, Wilcoxon rank-sum test) and 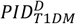 (p = 4 · 10^−9^, Wilcoxon rank-sum test). Similarly, in T2DM VPs, the optimal RL policy achieved the highest TIR, which was significantly greater than that achieved by both the *PID*_*T*2*DM*_ and *PI* 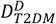 control algorithms (p = 3.8 · 10^−10^ and p = 1.7 · 10^−68^, Wilcoxon rank-sum test).

The TIR percentages obtained by the optimal RL policy *π*\* and the *PID* controller in both T1DM and T2DM VPs were strongly correlated (**Fig 4a**), being more noticeable in T1DM than in T2DM VPs. Conversely, the relationship between the optimal RL policy *π*\* and the *PID*^*D*^ controller exhibited a strong correlation for T1DM that was, however, almost negligible in T2DM VPs (**Fig 4b**). This enhanced performance of RL-based BG regulation was particularly evident in 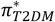, where the optimal RL policy achieved TIR values of 100% in most simulations. Taken together, the performance of the optimal RL policies *π*\* in achieving and maintaining normoglycemia exceeded the PID control algorithms.

As benchmarks of efficacy and safety, the percentages of TBR and TAR were evaluated. The optimal RL policy 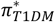 exhibited an almost negligible TBR of 0.02%, indicating a low risk of hypoglycemic events. In contrast, *PID*_*T*1*DM*_ and 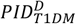 demonstrated significantly higher TBR percentages than 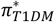 (p = 4.4 · 10^−50^ and p = 9.6 · 10^−39^, Wilcoxon rank-sum test). This finding is consistent with the lack of correlation observed in TBR percentages between 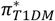 and *PID*_*T*1*DM*_ (**Fig 4c**), and 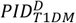 (**Fig 4d**). In T2DM VPs, the TBR achieved by 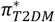 was significantly smaller than in *PID*_*T*2*DM*_ (p = 7.3 · 10^−28^, Wilcoxon rank-sum test) and in 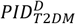 (p = 8.8 · 10^−76^, Wilcoxon rank-sum test). Due to the null variance of TBR values obtained by 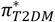, the calculation of the Pearson coefficients was not possible. Accordingly, a generalized functional discrepancy between the optimal RL policies and the *PID* or *PID*^*D*^ control algorithms in the management of hypoglycemia can be assumed.

In terms of hyperglycemia exposure, control algorithms for the T2DM VPs exhibited a considerably lower exposure than those for the T1DM VPs. However, differences in exposure between control algorithms within the same type of DM were less evident. In T1DM VPs, the optimal RL policy 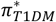 achieved the smallest TAR percentages, whereas differences to TAR percentages of *PID*_*T*1*DM*_ and 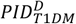 were not significant (see **Table 1**). Similarly, in T2DM VPs, all control algorithms could maintain low TAR percentages, and differences between the optimal RL policy 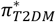 and the control algorithms *PID*_*T*2*DM*_ and 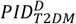 were not significant. Accordingly, a correlation was observed in the TAR percentages between the optimal RL policies *π*\* and the control algorithms *PID* (see **Fig 4e**) and *PID*^*D*^ (see **Fig 4f**) The findings suggest a high degree of functional coherence among all control algorithms for T1DM and T2DM under hyperglycemic conditions, with the algorithms for T1DM VPs once again exhibiting the strongest correlation.

The high correlation observed between the control algorithms in metrics such as TIR and TAR can be attributed to the existence of common patterns of response to analogous experimental conditions. Despite their design differences, both control algorithms similarly captured the underlying BG dynamics because of unforeseen disturbances, common physiological constraints, or a comparably constrained control variable. This suggests that, although the control with optimal RL policies showed better absolute performance, the behavior relative to experimental conditions was highly concordant, especially in T1DM VPs. Conversely, the absence of correlation of TBR percentages in T1DM and T2DM VPs suggests qualitative differences between control algorithms in managing hypoglycemia.

Taken together, all control algorithms were effective in regulating hyperglycemia, maintaining TAR percentages below the recommended threshold of 25% for the general population and 10% for older and/or high-risk individuals with DM [33]. These results emphasize the robust and safe performance of the optimal RL policies *π*\*, characterized by a minimal percentage of time spent in both hypo- and hyperglycemic ranges. However, potential safety concerns associated with *PID*^*D*^, particularly in relation to hypoglycemia in T2DM VPs, are indicated. Therefore, the optimal RL policies offer the most favorable balance between stability, efficacy, and safety under the conditions tested.

## Discussion

In this study, RL agents were trained to automatically maintain the BG concentration in the normoglycemic range. The resulting optimal RL policies *π*\* were tested by sampling patient-specific schedules of glucose loads. The optimal RL policies *π*\* were capable of determining the rate of adaptive insulin delivery required to fulfil the BG regulation targets set out in clinical guidelines, even in situations where disturbances are present, such as the administration of unannounced glucose loads or the presence of noise in CGM sensor measurements. The findings demonstrated that following the convergence of a maximum cumulative reward during the training phase, the resulting optimal RL policies *π*\* achieved a consistently high performance in both the training and validation environments. Simulations in the test environment showed that the optimal RL policies *π*\* could select insulin delivery rates that maximized the percentage of time during which BG remained within the normoglycemic range. Furthermore, the optimal RL policies *π*\* were able to prevent the occurrence of hypoglycemia in all simulations.

The optimal RL policies achieved notably higher TIR and lower TBR values compared to those reported in previous studies utilizing RL-based control algorithms. For instance, Zhu *et al*. implemented a basic hormone control approach to regulate BG in T1DM patients, resulting in a TIR of 80.9% and a TBR of 1.9% [25]. Likewise, Hettiarachchi *et al*. reported a TIR of 65.0% and a TBR of 3.1% when using an RL-based control strategy that required prior meal announcement [27]. These results contrast with the findings of the present study, as both studies report TIR lower than 97.3% and TBR values higher than 0.02%, achieved by the optimal RL policy 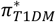. Further, the findings of this study contrast with those reported by Lim *et al*., whose SAC algorithm did not exhibit a significant performance advantage over a PID controller in any glycemic range in T1DM [29]. Similarly, they differ from the results of Fox *et al*., who observed only a 1% improvement in TIR over a PID controller when using the UVA-Padova simulator [34].

A comparison with the *PID* and *PID*^*D*^ control algorithms revealed superior performance of the optimal RL policies *π*\* in T1DM and T2DM patients, particularly indicated by a reduction of TIR and TBR percentages. Regarding TIR, a potential increase of 10%, as predicted for 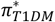 compared to 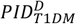, or of 15%, as predicted for 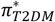 compared to 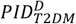, would represent a substantial therapeutic improvement. In this context, a previous study reported that an increase in TIR correlates with a reduction in glycosylated hemoglobin (HbA1c), a well-established clinical marker of long-term glycemic control that is strongly associated with diabetes-related morbidity and mortality. According to the study, a 10% increase in TIR was associated with an average reduction of 0.57% in HbA1c, while a 15% TIR increase corresponded to a reduction of 0.74% in HbA1c [35]. These findings underscore the clinical significance of even modest enhancements in TIR. Regarding TBR, it was found that only the optimal RL policies *π*\* fulfilled the target of maintaining TBR below 1% for older and/or high-risk diabetic patients [33], indicating that, while the PID strategies may provide effective BG control, they are associated with an elevated risk of hypoglycemia.

Although the outcome of the optimal RL policies *π*\* have been contrasted with the *PID* control algorithm, it is important to acknowledge that the latter is based on an idealised assumption of continuous BG measurements and analog insulin delivery (see **Table 4**). This indicates that the *PID* operates under the assumption of uninterrupted availability of CGM observations and infinitesimal fluctuations in the insulin delivery rate at any given time point. However, in practice, CGM systems measure interstitial BG at discrete intervals, typically every five minutes, and AID systems are restricted to the quantisation of the insulin delivery rate. It is therefore most realistic and representative to compare the performance of the optimal RL policies *π*\* with the *PID*^*D*^ control algorithm, as this model more accurately reflects the practical limitations inherent in current medical devices used in automated diabetes management.

While the present study offers promising results, it is important to acknowledge the limitations of the study design. Firstly, the *π*\* policies have been evaluated exclusively *in silico*, which limits the generalisability of the results to real patients. Furthermore, only oral glucose loads have been considered as a means of inducing fluctuations in BG. However, the potential influence of other dietary sources or more complex physiological situations, such as exercise or stress, on BG regulation in the daily lives of people with diabetes has not been considered. Notwithstanding these limitations, efforts have been made to ensure that the simulation environment accurately reflects the actual conditions to make it feasible in a real clinical setting. For instance, the sampling times of the system have been modified to correspond with the measurement intervals of commercially available CGM, and the insulin delivery ranges have been calibrated to align with the capabilities of current AID systems. Furthermore, CGM sensor noise has been simulated to approximate the inaccuracies of current devices, and the insulin reservoir capacity has been designed to mirror that of cartridges used in clinical practice.

As a prospective enhancement, the model could be expanded to encompass a dual hormone system, integrating both insulin and glucagon, or the conjunction of insulin with oral therapies such as metformin in T2DM patients. Furthermore, a pre-trained version of the optimal RL policy may be deployed in an AID. Subsequent to the acquisition of information derived from user usage, the AID could be capable of performing additional training to adjust the policy parameters based on individualised and personalised data. This would enable the policy to be adapted in accordance with individual responses to BG fluctuations, thereby facilitating more robust and adaptable BG regulation across a broader range of disturbances. Taken together, this work demonstrates a successful application of automated RL-based control algorithms for insulin administration in T1DM and T2DM, opening new possibilities for improved clinical management with minimal patient intervention.

## Methods

### RL-based blood glucose regulation

The closed-loop insulin delivery systems for BG regulation in DM can be conceptualized as interactions between an environment and an agent (see **Fig 1a**). The agent is linked to the delivery system and the control system, while the environment is associated with the BG dynamics observed through the measurements of a CGM over time. In the present study, for training and subsequent *in silico* testing, the glucose, insulin, glucagon and incretin dynamics of different organs and tissues of the body were described through a VP linked to a system of coupled ODEs. The environment dynamics governed by the set of ODEs are defined in continuous time, denoted by *τ* ∈ ℝ^+^. The interactions between agent and the environment were conducted in a series of discrete steps *t* = 0,1,2,…,*N* − 1. As part of a simulation, at the initiation of each step, the state of the system, designated as *s*_*t*_ ∈ *S*, was observed and transferred to the agent. The agent served as a decision-maker that, based on the state, performs an action *a*_*t*_ ∈ *A* ⊂ ℝ (*e*.*g*., selects the dose of insulin to be delivered). Actions were defined as decisions that affected the evolution of the system. Consequently, upon receiving an action, the environment evolved into a subsequent state, designated as *s*_*t*+1_. The function that defined the probability of undertaking an action based on the current state was referred to as a policy, expressed as *π* = *P*(*a*|*s*). The probability distribution *p*(*s*_*t*+1_|*s*_*t*_,*a*_*t*_) governed the transition between states and is dependent upon the current state and the action taken. The consequence of the action was assessed at the end of the step by a delayed reward, denoted by *r*_*t*+1_ ∈ *R* ⊂ ℝ. The reward quantified the efficacy of a specific action (*e.g*., achieving normoglycemic state) and was a function of the current state. Subsequently, the interaction started again with the following step until a complete episode was reached. Each episode *e* consisted of a sequence of states, actions and rewards, *e*.*g*., *e* = {*s*_0_, *a*_0_,*r*_1_,*s*_1_ *a*_1_,2…*s*_*N*−1_, *a*_*N*−1_,*r*_*N*_}.

The management of diabetes was formulated as a Markov Decision Process (MDP) problem represented by the tuple (*S*,*A*,*P*,*R*,*γ*), where *S* represented the set of possible states of the system, *A* is the set of possible actions, *R* was the reward function and *γ* ∈ [0,1] was the discount factor applied to future rewards. The objective of the agent was to learn the optimal RL policy *π*\* resulting in the maximization of the discounted accumulated total reward [17].

### Control systems perspective of blood glucose regulation in virtual patients

RL-based BG regulation can be described from a systems control engineering perspective (**Fig 1c**). Accordingly, the actions *a*_*t*_ of the agent served as the primary inputs to the environment, while the outputs consisted of the current state *s*_*t*_ and the corresponding delayed reward *r*_*t*+1_. At the core of this input-output transition, the VP linked variations in insulin delivery and food intake disturbances to the corresponding BG fluctuations, thereby influencing the reward signal.

Previous research has demonstrated the efficacy of implementing VP to train RL agents [25,27,34,36]. The VPs were constructed upon mathematical models which, when subjected to numerical simulation, predicted BG dynamics. Examples of mathematical models used for investigating insulin-glucose regulation include those developed by Cobelli and Kovatchev (UVA/Padova simulator) [37], Hovorka *et al*. [8], Ferdinando *et al*. [38], and Sorensen [39]. This research implemented the model developed by Sorensen [39], which is one of the most comprehensive and detailed mathematical models of DM pathophysiology that was expanded by Alvehag and Martin [40], and by López-Palau and Olais-Govea [41]. This model served to emulate the dynamics of BG in response to the homeostatic process or as a consequence of external factors, including oral glucose intake and exogenous insulin delivery (see **S1 Text** for details and T1DM and T2DM model equations).

The first input to the VP consisted of the subcutaneous insulin delivery rate *r*_*SCI*_ delivered by the close-loop system. It comprises two parts, the basal constant rate 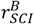, and an adaptive or variable rate 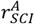:

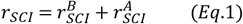

The basal constant rate 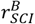 was designed to regulate fasting BG in the absence of endogenous insulin production. Therefore, it was considered as 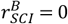 mU/min for T2DM patients and as 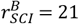 mU/min in T1DM patients as determined in the model by Sorensen [39]. The adaptive rate 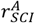 was intended to mostly regulate prandial BG. Therefore, the agent was capable of indirectly modulating BG through this input based on the state of the environment. The value of 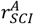 could be modified at the start of each step but remained unaltered throughout that step.

The rate of glucose entering the stomach *OGC*_*S*_, serving a as second input to a VP, was described by a trapezoidal pattern [40]

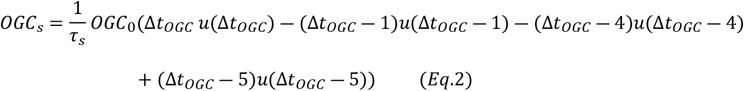

depending on the oral glucose load *OGC*_0_, the time delay Δ*t*_*OGC*_ = *τ* − *τ*_0_ in minutes between the simulation time and the time at which the oral glucose disturbance occurs *τ*_0_, a constant parameter *τ*_*s*_ associated with the time in minutes required for glucose ingestion, and the step function *u*(*τ*) representing the entry of glucose into the gut at the time *τ*. Based on *OGC*_*s*_, the rate of oral glucose absorption from the intestine to the blood was given by *r*_*OGA*_ [mg/dL/min] (see **S1 Text**, Eq. s24).

### Blood glucose concentration monitoring

The output of the VP was considered as the sampled blood glucose *BG*_*s*_ from the BG concentration in the vascular space of the periphery *G*_*PV*_(*τ*) (see **S1 Table**). Throughout the episodes, BG fluctuations were sampled in *T*_1_ min intervals to emulate the functionality of CGM devices in accordance with the technical specifications of the commercial CGM sensors due to the low dynamicity of the glucose-insulin system, *e*.*g*., *Dexcom G7* [12]. Values of the CGM sensor were defined as *BG*_*m*_(*t*_*p*_) = *BG*_*s*_(*t*_*p*_) + *n*. In this equation *t*_*p*_ is a discrete time with *p* set at 0 at the beginning of each episode and undergoes a sequential increment of one every *T*_1_ until the termination of the episode (i.e., *t*_*p*_ = *pT*_1_), and *n*~𝒩(0,*σ*^2^) was white noise of the CGM sensor with variance *σ*^2^. A value of *σ*^2^ = 3.677 was chosen based on the average variances observed in the *Dexcom G6* sensor by Vettoretti *et al*. [42]. After sampling, the measurements *BG*_*m*_ were maintained at a constant level between the measure intervals using a zero-order holder (ZOH). To gain insight into the rates of BG changes after glucose disturbances, the instant rate of change between the last two *BG*_*m*_ measurements of an episode was considered as the first observation variable. This rate of glucose change was computed as 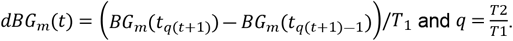.

To gain insight into the BG fluctuations between episodes, the average 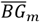, and an error function *BG*_*e*_ were included as the second and third components of the state. The average was computed as 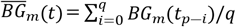. An error function of the form *BG*_*e*_(*t*) = *BG*_*m*_(*t*_*q*(*t*+1)_) − *BG*_*r*_ was calculated as the difference between the last BG measurements of an episode and a normoglycemic reference of *BG*_*r*_ = 110 mg/dL, which was also adopted as a reliable target for BG management in clinical guidelines [12].

The final state components incorporated a safety element designed to prevent hypoglycemic episodes. The attenuation factor *φ*(*t*) ∈ {0,1} was defined as:

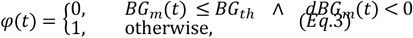

where a value of 0 results in the interruption of the insulin delivery. This value was achieved when *B G*_*m*_(*t*) reached or fell below a preset BG threshold *BG*_*th*_ = 100 mg/dL and a negative rate of glucose change was sustained. Conversely, a value of 1 indicated the restoration of adaptive insulin delivery. A comparable suspension mechanism has previously been implemented in commercial algorithms, for instance, the ‘prediction low glucose management’ suspension algorithm was integrated into the *Smart Guard* of the *Mini Med 640G* [12]. The set of system state elements was defined as 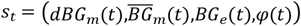, to provide information to the agent regarding the BG dynamics and the status of the insulin suspension mechanism.

Based on the state of the system, the agent selected the action necessary to bring and maintain the BG within the target range. Each action represented the constant rate of adaptive insulin to be delivered during the subsequent step. The set of permissible actions (*i*.*e*., action space *A*) was defined as a finite set of preset dosages: *A* = {*a*^1^,*a*^2^,…,*a*^16^} with constant increments from 0 to *I*_*max*_. This is *a*^*j*^ = *I*_*max*_ (*j* − 1)/15, where *I*_*max*_ = 150 mU/min. Thereby, *a*^1^ corresponded to no adaptive or variable rate of insulin to be given regarding *φ*, therefore, minimal dose was set to be 0 mU/min. The selection of an RL control scheme resulted in the formulation of an adaptive insulin delivery scheme with the form 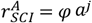.

The upper limit for insulin delivery was set at 300 U representing the maximum reservoir capacity. Before releasing the drug, the amount of drug available at each step was verified. If the dose was less than the amount of drug available, the requested amount was released. On the other hand, if the available amount of drug was less than the dose, then the available amount was released until it was depleted. Once the action was selected and the adaptive insulin delivery rate was calculated, it was transmitted to the environment, specifically to the VP model, and remained constant throughout the entire step. The action space was identical for both BG regulation in T1DM and T2DM patients.

### Reward function for blood glucose regulation

The reward function, as a key element in the decision-making process of the agent, was used to guide its actions with the objective of delivering insulin. The reward function *R* constituted a heuristically defined mapping that transferred the measured BG into numerical values *R*:(*s*_*t*_,*a*_*t*_)→*r*. It was defined as the sum of a set of functions depending on state variables and have specific objectives. The following objectives were translated to components of the reward function *R*_1_, *R*_2_ and *R*_3_:

(1) To maintain BG within the normoglycemic range (*i*.*e*., 90…120 mg/dL). This objective was accomplished through the implementation of a consistent reward structure dependent on the maintenance of mean BG within the normoglycemic range

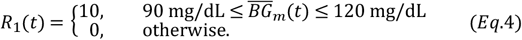

(2) To promote rapid convergence to normoglycemic state. To achieve this objective, it was necessary to consider the value of the reward as a function of the BG rate of change in combination with the glycemic state

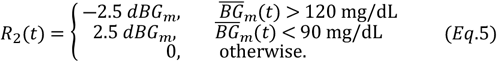

(3) To highly penalize severe hypoglycemia. In instances where the BG concentration fell below 70 mg/dL, a significant penalty was imposed due to the proximity of the BG to the threshold of hypoglycemia

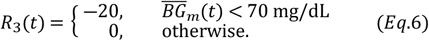

Taking together the parts of Eqs. 4–6, the reward function was defined as *r*_*t*_ = *R*_1_(*t*) + *R*_2_(*t*) + *R*_3_(*t*). By evaluating the reward function at the end of a step, the obtained reward *r* was a unitless scalar value providing feedback to the agent, thereby enabling it to learn to maintain BG within the normoglycemic range and to enhance the policy. A schematic of the objective function can be seen in **Fig 1b**. The maximum reward was obtained in the normoglycemic range between 90 and 120 mg/dL.

### Implementation of the actor and critic artificial neural network

A stochastic single agent with an actor-critic structure was implemented for optimizing the regulation of BG dynamics. The actor *π*(*a*_*t*_|*s*_*t*_;*θ*) with parameters *θ* delivered the conditional probability of taking each action *a*_*t*_ when in state *s*_*t*_ at time *t*. The critic *V*(*s*_*t*_;*ϕ*) with parameters *ϕ*, related the observation *s*_*t*_ to the corresponding expectation of the discounted long-term reward [43]. The objective of the actor was to learn the policy by obtaining feedback from the critic. Conversely, the objective of the critic was to learn the value function, as described below that was implemented for determining the goodness of a given state. The actor and the critic were function approximators parameterized with artificial neural networks (ANN).

A detailed description of the architecture of the networks of actors and critics is given in **Table 2**. The actor and the critic comprised of 8- and 7-layer ANNs, respectively. Both ANN were fed by a sequence input layer of 4 nodes corresponding to the elements of the state of the environment. The ANNs consisted of fully connected layers, Rectified Linear Unit (ReLU) activations and a layer with a Gated Recurrent Unit Layer (GRU) of 350 nodes to give recurrence to the network and learn dependencies between the data of the different steps. The actor was represented by a state-action function. Accordingly, the output layer was a SoftMax layer with 16 nodes representing possible actions. The critic is a state-value function. Therefore, the output layer contained a single node representing the value of the future reward. During the training phase, the parameters *θ* and *ϕ* were updated using a gradient descent algorithm.

**Table 2.** Architectural design of the actor-critic artificial neural networks.

|  |  |  |  |  |
| --- | --- | --- | --- | --- |
| Layer | **Actor ANN** |  | **Critic ANN** |  |
|  | Type | Activations* | Type | Activations* |
| 1<sup>st</sup> | Sequence Input | $4(C) \times 1(B) \times 1(T)$ | Sequence Input | $4(C) \times 1(B) \times 1(T)$ |
| 2<sup>nd</sup> | Fully Connected | $700(C) \times 1(B) \times 1(T)$ | Fully Connected | $700(C) \times 1(B) \times 1(T)$ |
| 3<sup>th</sup> | ReLU | $700(C) \times 1(B) \times 1(T)$ | ReLU | $700(C) \times 1(B) \times 1(T)$ |
| 4<sup>th</sup> | Fully Connected | $700(C) \times 1(B) \times 1(T)$ | Fully Connected | $700(C) \times 1(B) \times 1(T)$ |
| 5<sup>th</sup> | ReLU | $700(C) \times 1(B) \times 1(T)$ | ReLU | $700(C) \times 1(B) \times 1(T)$ |
| 6<sup>th</sup> | GRU | $350(C) \times 1(B) \times 1(T)$ | GRU | $350(C) \times 1(B) \times 1(T)$ |
| 7<sup>th</sup> | Fully Connected | $16(C) \times 1(B) \times 1(T)$ | Fully Connected | $1(C) \times 1(B) \times 1(T)$ |
| 8<sup>th</sup> | SoftMax | $16(C) \times 1(B) \times 1(T)$ | | |
| * Labels of the activation dimension: (C) Channel, (B) Batch observations, (T) Time or sequence |  |  |  |  |

### Training process

The initial condition for each simulation episode was defined as the basal postabsorptive state of the VP. In the basal state, the time derivatives of all model state variables equaled zero. Concurrently, the metabolic rates assumed their basal values, thereby achieving effective decoupling of the glucose, insulin, glucagon and incretin subsystems. Consequently, by setting a single BG concentration (for both T1DM and T2DM VPs) and a single insulin concentration (for the T2DM VPs), all other state variables at steady state were uniquely determined. For simplicity, the BG and insulin concentrations used to calculate state variables at steady state were selected by randomly sampling basal peripheral venous concentrations from uniform distributions. For T1DM VPs, the distribution was determined as *U*[106.74 mg/dL, 279.71 mg/dL] using the values reported by Zhao, *et al*. [44], while for T2DM was determined as *U*[168 mg/dL, 192 mg/dL] and *U*[16.6 mU/L, 22.6 mU/L] using the values reported by Abdul-Ghani, *et al*. [45]. The resulting steady-state variables were used to define the basal concentrations embedded within specific model parameters (see **S6 Table**) and served as initial conditions during the first numerical integration step of each simulation. The integration of the model was conducted using the MATLAB function ODE15S.

A cohort of 2000 VPs with T1DM and 2000 with T2DM was generated by specifying distinct basal postabsorptive states. Three quarters of these patients were used for training and the remaining quarter for validation. During the episodes, in each VP, one oral glucose intake was simulated. The glucose load *OGC*_0_ and the time of ingestion *τ*_0_, were defined as scalar values arbitrarily selected from the uniform distributions *OGC*_0_~*U*[0 g,80 g] and *τ*_0_~*U*[0 min,180 min] min.

The PPO algorithm, developed by Schulman *et al*. [46], was used to estimate probabilities of taking each action in the action space, randomly selecting actions based on the probability distribution and updating the ANN parameters during training. **Fig 5** shows a schematic of the agent training using the PPO agent ‘rlPPOAgent’ from MATLAB [47]. Initially, the actor and critic ANNs were defined with arbitrary parameters *θ*_0_ and *ϕ*_0_, respectively. Employing the initial policy *π*_0_, a total of *E* experiences were generated and stored in the form {*s*_*t*_, *a*_*t*_,*r*_*t*+1_,*s*_*t*+1_}. The *E* experiences were divided into sequences of *N*_*E*_ experiences, numbered from *ts* = 1 at the beginning of a training episode, to *ts* + *N*_*E*_ with *N*_*E*_ as the experience horizon. For each step *t* = *ts, ts* + 1,…,*ts* + *N*_*E*_ −1 of each sequence, the advantage function *D*_*t*_, and the return function *G*_*t*_ were computed and stored. The advantage function, computed as *D*_*t*_ = *G*_*t*_ −*V*(*s*_*t*_;*ϕ*), represents a measurement of the performance of the selected action compared to the average of the possible actions under state *s*_*t*_. The return function represents the discounted reward accumulated over time. This function is computed as 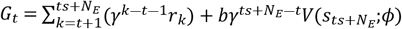, where *b* is a parameter that equals 0 if 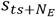 is a terminal state and 1 otherwise, and *γ* = 0.99 equally considered short-term and long-term rewards.

**Fig 5.**
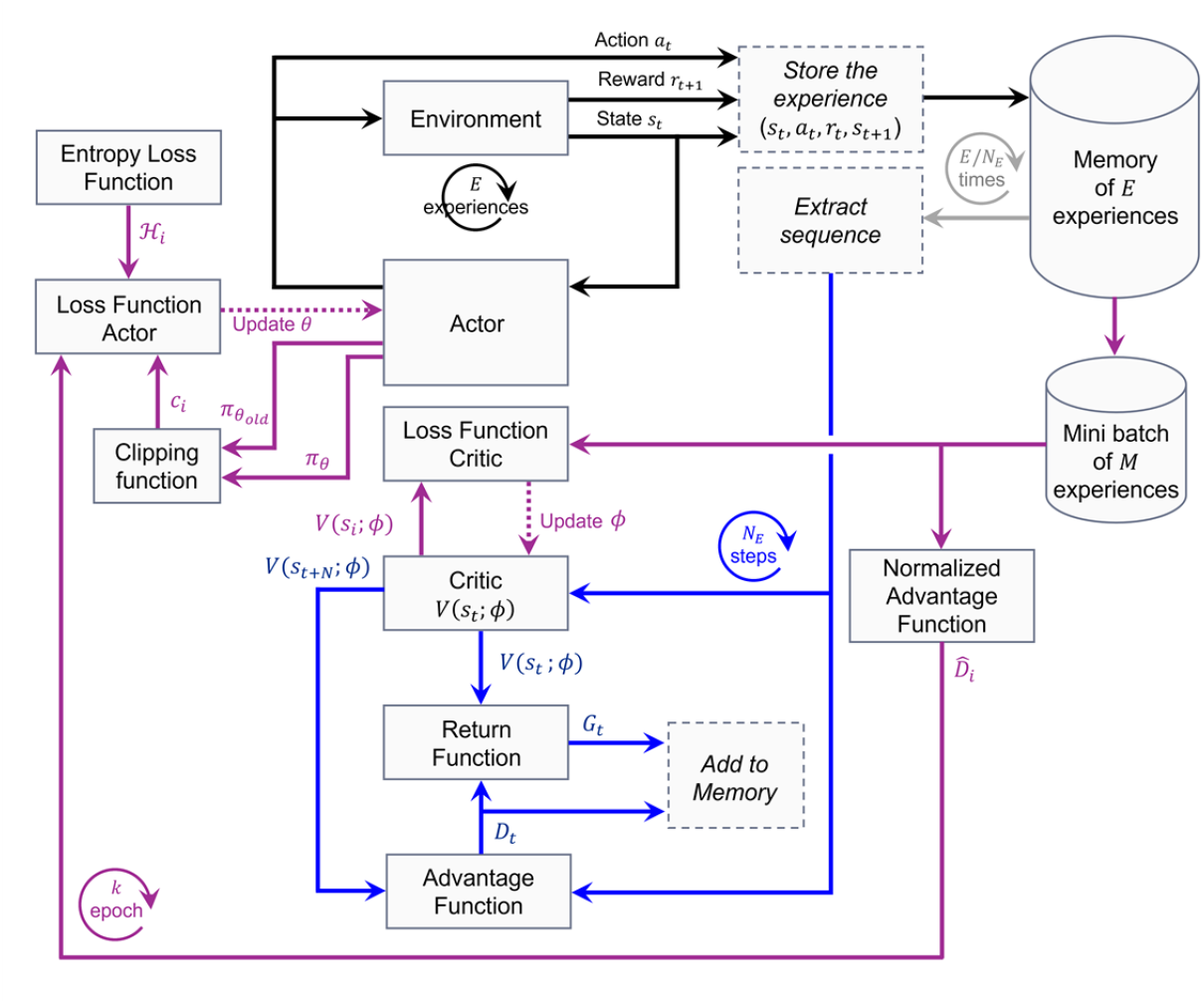
Block diagram of the learning process using the PPO algorithm. The arrows represent the progression of information throughout the learning process. The black arrows represent the initial cycle of data collection and storage, in which *E* experiences are accumulated and retained within the buffer. The grey lines represent the process by which the total number of *E* experiences were extracted in sequences of *N*_*E*_ experiences. For each step in each sequence of *N*_*E*_ experiences, the advantage and return functions were calculated (*i*.*e*., blue lines cycle). In the final stage of the process, the neural networks of the critic and the actor are adapted each *k* epoch by minimizing the loss functions calculated from a set of *M* preselected experiences in each mini-batch (*i*.*e*., magenta cycle).

Along *k* = 7 epochs, the set of *E* experiences was divided into *B* = 100 mini batch of *M* = 24 selected experiences. The information contained in each batch was used to update the parameters of the critic and the actor ANN. The parameters of the ANN of the critic were updated by iteratively minimizing the loss function of the critic, defined as 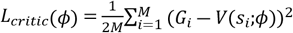. Accordingly, the parameters of the actor ANN were updated by minimizing the loss function of the actor, computed as 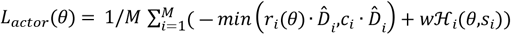, were 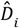 represent the normalized advantage values defined by 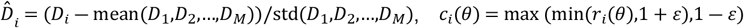 was the clipping function with *r*_*i*_(*θ*) = *π a*_*i*_|*s*_*i*_;*θ* /*π a*_*i*_|*s*_*i*_;*θ*_*old*_ as the probability ratio 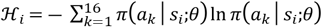 was the entropy function related to exploration of the environmental space by the agent, and *w* was the entropy loss weight [47]. In order to encourage exploration and enhance the stability of the learning process, an entropy loss weight *w* = 0.7 and a clip factor *ε* = 0.3 were selected for both agents. Furthermore, to prevent the premature convergence and exploration of the action space the learning rate of the actor and critic was tuned in to both agents. **Table 3** presents an overview of the hyperparameters implemented.

**Table 3.** Hyperparameters for agent training and deployment.

|  | Type 1 diabetes mellitus |  | Type 2 diabetes mellitus |  |
| --- | --- | --- | --- | --- |
| Clip factor ($\epsilon$) | 0.3 | | 0.3 | |
| Entropy loss weight ($w$) | 0.7 | | 0.7 | |
| Discount factor ($\gamma$) | 0.99 | | 0.99 | |
| Experience horizon ($N_E$) | 180 | | 240 | |
| Number of epochs ($k$) | 7 | | 7 | |
| Mini batch size ($M$) | 24 | | 24 | |
| Learning frequency ($E$) | $10 \cdot M$ | | $10 \cdot M$ | |
| Advantage estimation method | Finite-horizon |  | Finite-horizon |  |
| Advantage normalization method | current |  | current |  |
|  | **Actor** | **Critic** | **Actor** | **Critic** |
| Learning rate | $3.8 \cdot 10^{-5}$ | $4.4 \cdot 10^{-4}$ | $2.5 \cdot 10^{-5}$ | $4.5 \cdot 10^{-5}$ |
| Gradient threshold | Inf | Inf | Inf | Inf |
| Optimizer | adam | adam | adam | adam |
| Gradient decay | 0.9 | 0.9 | 0.9 | 0.9 |
| Gradient threshold method | l2norm | l2norm | l2norm | l2norm |
| L2 regularization | $1 \cdot 10^{-4}$ | $1 \cdot 10^{-4}$ | $1 \cdot 10^{-4}$ | $1 \cdot 10^{-4}$ |

As the training iterations progressed, there was a discernible improvement in the performance of the agents, as evidenced by the increasing average reward. The training was concluded by the stable convergence of the discounted cumulative average reward over a 10-episode window to a maximum value.

### Validation and testing of optimal RL policies

Upon completion of the training phase, simulations were conducted in the validation environment. Results from this simulation were compared to the results of training to assess the efficacy of the optimal RL policy in an unseen dataset. The efficacy of the optimal RL policy to generalize in a more representative setting was assessed by simulations conducted in a test environment. The test environment was designed to evaluate the performance of the optimal RL policy in more generalized situations. In this environment, no further learning or policy adjustments were conducted. Each episode comprised *N* = 96 steps (equivalent to 24 hr.). Both *OGC*_0_ and *t*_0_ were three-element vectors randomly selected from the uniform distributions 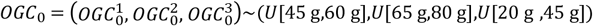 and 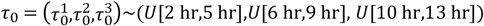. For the purposes of testing, n=200 VPs were created to interact with the optimal RL policy within the designated test environment. It was determined that these patients exhibited three oral disturbances on the same day. The glucose load and time of intake were randomly selected from the aforementioned uniform distributions.

Based on the control BG targets for CGM measurements, as consented in the *Advanced Technologies & Treatments for Diabetes 2019* [32], the mean BG concentration, the coefficient of variation of BG, and percentages of time within BG ranges (TIR, within the normoglycemic range; TBR, below the normoglycemic range; TAR, above the normoglycemic range) were implemented as measures of performance. To assess the dispersion, the mean BG values 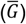 and the coefficients of BG variation (CV) were calculated. The coefficient of BG variation was defined as 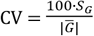, with *S*_*G*_ being the standard deviation of BG concentration. The time in range (TIR) was defined as the percentage of the episode time when the BG concentration was between 70 and 180 mg/dL [48]. Accordingly, TBR and TAR were denoted as percentages of the episode time when BG was below 70 mg/dL and above 180 mg/dL, respectively. The primary goal was to increase the percentage of TIR while reducing the TBR and TAR, to avoid chronic microvascular and macrovascular complications [33], and prevent acute events resulting from severe BG dysregulation potentially resulting in loss of consciousness and death.

**Table 4** shows a list of the control algorithms used along the training, validation and testing process. Simulations were compared to the performance of a PID initialized on the same fasting BG and subjected to the same disturbances. A detailed description of the PID equations and tuning is provided in **S2 Text**. Specifically, **S1 Fig** illustrates the implemented block diagram of the closed-loop PID control algorithm for BG regulation, whereas **S7 Table** summarizes the parameter values employed in the PID control implementation.

**Table 4.** Control algorithms description and features.

| Table 4. Control algorithms description and features. |  |  |  |  |  |  |  |  |
| --- | --- | --- | --- | --- | --- | --- | --- | --- |
| Control algorithm | VP type |  | Observations |  | Control signal |  | Trained |  |
|  | T1DM | T2DM | Discrete | Continuous | Digital | Analog | Yes | No |
| $\pi_{T1DM}^0$ | ■ | | ■ | | ■ | | | ■ |
| $\pi_{T2DM}^0$ | | ■ | ■ | | ■ | | | ■ |
| $\pi_{T1DM}^*$ | ■ | | ■ | | ■ | | ■ | |
| $\pi_{T2DM}^*$ | | ■ | ■ | | ■ | | ■ | |
| $PID_{T1DM}$ | ■ | | | ■ | | ■ | - | - |
| $PID_{T2DM}$ | | ■ | | ■ | | ■ | - | - |
| $PID_{T1DM}^D$ | ■ | | ■ | | ■ | | - | - |
| $PID_{T2DM}^D$ | | ■ | ■ | | ■ | | - | - |
| - | Not applicable |  |  |  |  |  |  |  |

## Acknowledgements

Not applicable.

## Code Availability

The underlying code for this study is not publicly available but may be made available to qualified researchers on reasonable requests from the corresponding author.

## Data Availability

All data generated or analyzed during this study are included in this published article and its Supporting Information files.

## Author contributions

NELP contributed to the conceptualization of the problem, participated partially in programming, conducted the training process, analyzed the results, and contributed to writing the manuscript. PNM contributed to discussions on methodology and assisted with manuscript revisions. JS contributed to discussions concerning the theoretical foundations of the problem. RE contributed to discussions concerning the theoretical foundations of the problem and assured funding. SMK contributed to the conceptualization of the problem, participated partially in programming and writing the manuscript. All authors read and approved the final version of the manuscript.

## Human Ethics and Consent to Participate declarations

Not applicable.

## Supporting information captions

**S1 Text. Mathematical model**. The present text offers a comprehensive overview of the compartmental mathematical models that have been developed to represent the pathophysiology of type 1 and type 2 diabetes mellitus. These models, formulated through systems of differential equations, describe blood glucose dynamics in response to oral glucose disturbances in the virtual patients. The text comprises **S1**–**S4 Tables**, which detail the equations corresponding to the glucose, insulin, glucagon, and incretin subsystems, respectively. Furthermore, **S5 Table** contains the auxiliary equations employed in the models, and **S6 Table** provides a description of the parameters implemented in each model.

**S2 Text. PID tuning**. This text provides a brief overview of the fundamental principles of PID control and a detailed description of the tuning methods used for both continuous and discrete PID controllers (*i*.*e*., *PID* and *PID*^*D*^). In this text, the **S1 Fig** shows the closed-loop control block diagram for the glucose regulation process in patients with diabetes. Additionally, **S7 Table** reports the parameter values used by the PID controllers after applying the Ziegler–Nichols tuning method. The resulting PID controllers were implemented as reference methods to facilitate a comparative analysis of the performance of the RL optimal polices *π*\*.

